# Electroconvulsive therapy effects on anhedonia and reward circuitry anatomy: a dimensional structural neuroimaging approach

**DOI:** 10.1101/2021.11.16.21266190

**Authors:** Marta Cano, Erik Lee, Alexis Worthley, Kristen Ellard, Tracy Barbour, Carles Soriano-Mas, Joan A. Camprodon

**Author notes:** **Corresponding author.** Carles Soriano-Mas, PhD, Department of Psychiatry, Bellvitge University Hospital-IDIBELL, s/n Feixa Llarga, Hospitalet de Llobregat, 08907, Barcelona, Spain. **Corresponding author.** Joan A. Camprodon, MD, PhD, Department of Psychiatry, Massachusetts General Hospital, Harvard Medical School, 149 13^th^ Street, Charlestown, 02129, Massachusetts, USA.

## Abstract

Anhedonia is a core symptom of major depressive disorder (MDD) resulting from maladaptive reward processing. Electroconvulsive therapy (ECT) appears to be an effective treatment for patients with treatment-resistant depression (TRD). However, no previous neuroimaging studies have taken a dimensional approach to assess whether ECT-induced gray matter (GM) volume changes are specifically related to improvements in anhedonia and positive valence emotional constructs. Here we aimed to assess the relationship between right unilateral (RUL) ECT-induced brain volumetric changes and improvement in anhedonia and reward processing in patients with TRD. We evaluated 15 patients at two time points (before the first ECT session and after acute ECT completion) using magnetic resonance imaging (MRI), clinical scales (i.e., Quick Inventory of Depressive Symptomatology [QIDS] for syndromal depression severity and Snaith-Hamilton Pleasure Scale [SHAPS] for anhedonia) and the Temporal Experience of Pleasure Scale (TEPS) for anticipatory and consummatory experiences of pleasure. Patients with TRD showed a significant improvement in anhedonia symptoms and both anticipatory and consummatory pleasure after RUL ECT completion. Moreover, GM volume increases within the right reward system were related to anhedonia responders and, specifically, improvement in anticipatory (but not consummatory) reward. We highlight the importance of a dimensional and circuit-based approach to understanding target engagement and the mechanism of action of ECT, with the goal to define symptom- and circuit-specific response biomarkers for device neuromodulation therapies.

## INTRODUCTION

Electroconvulsive therapy (ECT) remains the most effective treatment in psychiatry, and among the most effective in medicine. It is indicated primarily for major depressive disorder (MDD), bipolar depression (BD) and mania (BM), schizophrenia (SZ) and catatonia [1,2]. In the case of MDD, response rates are in the 70-90% range, which is remarkable given how severe and treatment-resistant ECT patients are [3].

While our clinical understanding of ECT is significant, it contrasts with how little we know about its neurobiological mechanisms of action. Given how diffuse the induced electric fields are [4] and the fact that ECT leads to a generalized seizure (i.e., the entire brain seizes), ECT has been traditionally considered a diffuse and non-specific neuromodulation treatment [5,6]. However, previous neuroimaging ECT research has shown ECT is more focal than previously considered [7,8].

A series of clinical observations also challenge this non-focal assumption and have motivated our current research. At the syndromal level, ECT is indicated for the conditions mentioned above, but it is not used to treat all neuropsychiatric disorders [1]. This suggests that while the entire brain seizes, only the circuits maladaptively affected by MDD, BD/BM and SZ change in a therapeutic direction, while the brain structures pathologically compromised in other conditions such as anxiety disorders, obsessive-compulsive disorder or eating disorders (for example) do not seem to be therapeutically modulated, providing further evidence for ECT specificity and focality.

Another line of evidence comes from a dimensional analysis of the effects of ECT. Certain domains such as reward processing seem to be maladaptive in the different clinical syndromes for which ECT is indicated (e.g., anhedonia in MDD or BD, or negative symptoms in SZ) and respond to ECT in a transdiagnostic manner [9]. Beyond the therapeutic benefits, a dimensional analysis of the iatrogenic effects of ECT also suggests focality: while ECT can particularly worsen memory [10,11], it does not affect other cognitive domains such as language, and other functions such as spatial attention and executive function seem to also be preserved or minimally affected (except in the cases of post-ictal delirium) [12,13]. Therefore, one can hypothesize that ECT may lead to relatively focal and specific modulation of reward and memory circuits, while it spares language, spatial attention or executive function networks.

Building from these clinical observations we took a dimensional and circuit-focused approach to study the mechanism of action of ECT, aiming to understand its impact on reward processing (clinical) and reward circuitry structure (anatomy). Reward processing represents a key positive valence affective and behavioral dimension of transdiagnostic pathological relevance [14]. There is growing recognition that anhedonia does not represent a unitary dimension; among its subcategories, two constructs emerge with clear relevance to behavior and disease: reward anticipation and consummation [15–17].

In this study we assessed patients with treatment-resistant depression (TRD) undergoing ECT before and after the acute course of treatment using magnetic resonance imaging (MRI), clinical and dimensional psychometric tools assessing depression severity, as well as anhedonia and its dimensional subconstructs (anticipation and consumption). Our aims were to understand the impact of ECT (1) on *clinical and dimensional constructs* of pathological reward processing, (2) the *anatomical* effects on core nodes of the reward circuitry, and (3) whether the anatomical changes in this circuitry were associated with the putative clinical improvement in anhedonia and its subconstructs.

## MATERIALS AND METHODS

### Participants

Fifteen patients with TRD were recruited from the Massachusetts General Hospital ECT service. Psychiatric diagnoses were established using clinical interview and the Mini-International Neuropsychiatric Interview 6.0 (MINI) [18]. All patients underwent a clinical assessment using the Quick Inventory of Depressive Symptomatology (QIDS) [19] for disorder severity and the Snaith-Hamilton Pleasure Scale (SHAPS) [20] for anhedonic symptomatology. Clinical response was defined as a decrease of 50% or more on QIDS and/or SHAPS scores. Moreover, the Temporal Experience of Pleasure Scale (TEPS) [21] was administered to evaluate anticipatory and consummatory subconstructs of reward processing. Exclusion criteria included: (i) the presence or past history of a severe medical or neurological disorder, (ii) contraindication to magnetic resonance imaging (MRI) scanning or abnormal MRI upon visual inspection, and (iii) a history of ECT during the past 12 months. Pharmacological treatment was maintained unchanged throughout the ECT protocol. The study was approved by the local ethics committee and was performed in compliance with the Declaration of Helsinki. All participants gave written informed consent to participate in this study.

### Electroconvulsive therapy

The fifteen patients were treated with ultra-brief and brief pulse (0.3-0.5 ms) ECT using a MECTA spECTrum 5000Q machine (Portland, Oregon). All patients started treatment with right unilateral (RUL) ECT (D’Elia electrode placement [22]), but three transitioned to bilateral (BIL) treatment given poor initial response (8 RUL + 12 BIL ECT sessions in one patient, 11 RUL + 10 BIL ECT sessions in other patient and 10 RUL + 5 BIL ECT sessions in the other). Anesthesia for the procedure was provided using methohexital (0.8-1.2 mg kg^-1^) for induction and succinylcholine for muscle relaxation (0.5-1 mg kg^-1^). Initial stimulus dose was determined by titrating the seizure threshold, starting with 19 millicoulombs for women and 38 millicoulombs for men. Subsequent treatments were performed at six times the estimated seizure threshold. During the acute course of ECT, treatments occur thrice a week every other day. The number of treatments range was 6-21 (mean = 12.07).

### Image acquisition and preprocessing

All the patients were scanned two times: before the first ECT session (MRI1) and after completion of the acute ECT course (MRI2). A 3.0-T Siemens Skyra scanner (Munich, Germany) equipped with a 32-channel head coil was used. A total of 156 slices were acquired with repetition time = 2530 ms; multi-echo time = 1.69, 3.55, 5.41, 7.27 ms; flip angle = 7°; field of view = 256×256 mm; matrix size 256×256 pixels; in-plane resolution = 1 × 1 mm^2^; slice thickness = 1 mm.

Structural MRI data were processed on a Microsoft Windows platform using technical computing software (MATLAB 7.14; The MathWorks, Natick, MA, USA) and Statistical Parametric Mapping (SPM12; The Welcome Department of Imaging Neuroscience, London, UK). The preprocessing consisted of an initial rigid-body within-subject coregistration to the first scan (MRI1) to ensure good starting estimates. This was followed by a pairwise longitudinal registration between the scans of each participant to obtain an average image and a Jacobian difference map. The average image was segmented, and the GM voxels were multiplied by the Jacobian difference map to obtain a GM volume change map for each participant. Next, we generated one specific template of our study sample (in Montreal Neurological Institute (MNI) space) using a Diffeomorphic Anatomical Registration, which was used to spatially normalize the GM volume change maps. Finally, images were smoothed with a 6 mm full-width at half maximum isotropic Gaussian kernel.

### Region of Interest (ROI): selection and justification

In order to assess structure-symptom relationships within the reward system we defined five anatomical masks comprising regions of interest (ROIs) in the reward network: nucleus accumbens (NAcc), ventral tegmental area (VTA), hippocampus, amygdala, and medial orbitofrontal cortex (mOFC). Nucleus accumbens-ventral tegmental area (NAcc-VTA) projections via the medial forebrain bundle have been consistently characterized as the central pathway of the reward circuitry [23]. Notwithstanding, the baseline state of NAcc-VTA circuit is known to be modulated by hippocampal activity, positioning the hippocampus as an important reward pacemaker into the hippocampus-NAcc-VTA loop [24]. In addition, the amygdala has a critical role in emotional coding of negative and also positive valence emotional stimuli, and amygdala-NAcc interactions are implicated in reward processing [25]. Finally, the NAcc-VTA pathway receives its main cortical input from the mOFC, and these interactions have been highlighted as a potential mechanism of cognitive reward representation [26]. Importantly, an additional ROI implicated in depression and mood but not canonically considered part of the reward system, the Brodmann area 25 (BA25), was included as an anatomical control [27]. We selected right-hemisphere structures as these patients received right unilateral ECT and previous research (and our present results) have shown that the structural effects of ECT are lateralized to the hemisphere being stimulated [8]. NAcc, amygdala, hippocampus, mOFC and BA25 masks were created using the WFU_PickAtlas toolbox (Wake Forest University School of Medicine) as implemented in SPM12 [28]. For the VTA, we used the mask defined by Murty et al., [29], which was automatically divided into right and left parts using SPM12.

### Statistical analyses

Sociodemographic, clinical and reward-related psychometric data were analyzed with SPSS version 21 (SPSS, Chicago, IL, USA) using nonparametric tests (Table S1).

#### Neuroimaging within-group analysis

To assess RUL ECT effects on GM volume changes, we performed a whole-brain one-sample model. Age and gender were included as confounding covariates. Statistical significance was set at *p*<0.05, family-wise error corrected (FWE) for whole-brain multiple comparisons.

#### Relationship with clinical response

The association between clinical response and GM volume changes was assessed using two independent multiple regression analyses (to assess continuous relationships) and 2 two-sample *t*-test models (responders versus non-responders), resulting from the independent analysis of the pre-post treatment change in two clinical scores (QIDS and SHAPS). Age and gender were included as confounding covariates in all analyses. These analyses were restricted to the six ROIs described above, and, for each ROI, statistical significance was set at *p*<0.05, FWE corrected for multiple comparisons across all in-mask voxels (i.e., using small-volume correction procedures). Nevertheless, significance was further adjusted using false discovery rate (FDR) to account for the six independent comparisons [30].

#### Relationship with anticipatory and consummatory subconstructs

The association between pre-post treatment anticipatory and consummatory pleasure changes and GM volume changes was evaluated using two independent multiple regression analyses, with age and gender as confounding covariates. As above, these analyses were restricted to the six ROIs, and, within each ROI, statistical significance was set at *p*<0.05, FWE corrected for multiple comparisons across all in-mask voxels, while FDR correction was used to account for the total number of comparisons.

## RESULTS

### Clinical and dimensional response

The mean ± s.d. QIDS score prior to ECT initiation was 18±3.38 and the mean QIDS ± s.d. score after the completion of the ECT was 12±5.21. The reduction in depression severity (30.40%, as measured by the percentage of change in QIDS score; 33.34% response rate) was significant according to a Wilcoxon signed-rank test (*z*=-2.844; *p*=0.004). In addition, the mean ± s.d. SHAPS score prior to ECT initiation was 5.07±2.76 and the mean SHAPS ± s.d. score after the completion of the ECT was 2.47±3.31. The improvement in anhedonia (52.16%, as measured by the percentage of change in SHAPS score; 66.67% of response rate) was significant according to Wilcoxon signed-rank tests (*z*=-2.744; *p*=0.006).

The mean ± s.d. TEPS scores prior to ECT initiation were 41±9.91 for the anticipatory score and 42.20±9.99 for the consummatory score and the mean TEPS ± s.d. scores after the completion of the ECT were 50.47±9.02 for the anticipatory score and 45.67±9.19 for the consummatory score. The improvements in both dimensional reward constructs (17.96%, as measured by the percentage of change in anticipatory TEPS score, and 7.88% as measured by the percentage of change in consummatory TEPS score) were significant according to Wilcoxon signed-rank tests (anticipatory TEPS, *z*=-2.983; *p*=0.003; consummatory TEPS, *z*=-2.705; *p*=0.007). In addition, the percentage of change in the anticipatory TEPS score was significantly greater than the percentage of change in the consummatory TEPS score (17.96% vs. 7.88%; Wilcoxon signed-rank test, *z*=-2.669; *p*=0.008).

### Neuroimaging within-group analysis

Patients with TRD receiving ECT showed volume increases encompassing right hemisphere regions including the striatum (i.e., NAcc, putamen and pallidum), the medial temporal lobe (i.e., amygdala and hippocampus), the anterior insular cortex, the anterior midbrain (substantia nigra/ventral tegmental area, SN/VTA), the subgenual anterior cingulate cortex (SgACC), and the perigenual anterior cingulate cortex (PgACC). These results are summarized in Table S2 and Figure 1.

**Figure 1.**
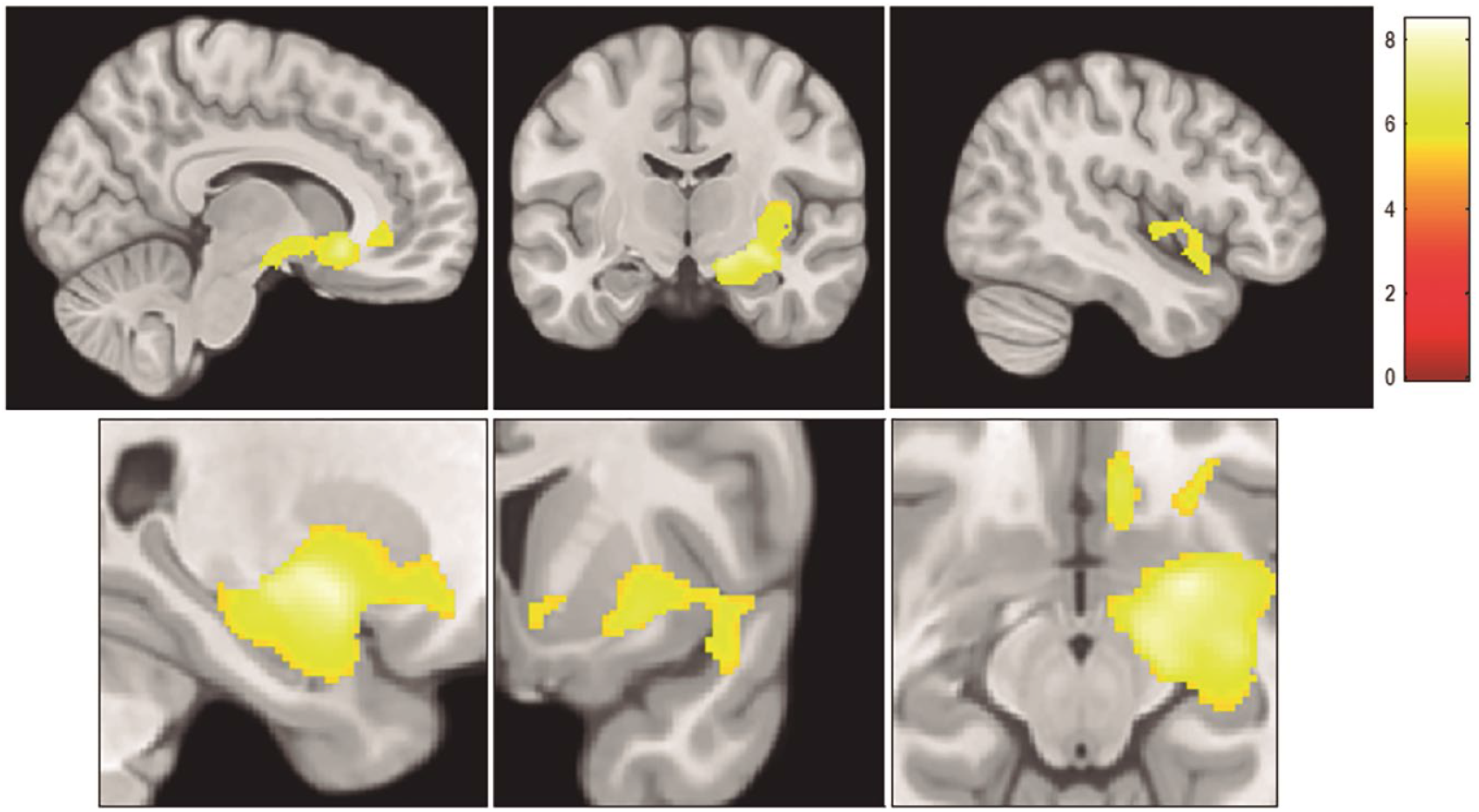
Clusters of volume increase, overlaid onto an MNI-normalized brain template, encompassing right hemisphere regions including the striatum (i.e., nucleus accumbens (NAcc), putamen and pallidum), the medial temporal lobe (i.e., amygdala and hippocampus), the anterior insular cortex, the anterior midbrain (substantia nigra/ventral tegmental area, SN/VTA), the subgenual anterior cingulate cortex (SgACC), and the perigenual anterior cingulate cortex (PgACC). For representation purposes, results are displayed at a significance threshold of *p*<0.0001 uncorrected at the voxel level and a cluster extent threshold of 1392 voxels. Left hemisphere is depicted on the left. Color bar represents *t*-value.

### Relationship between structural changes and clinical response

When analyzing the correlation between clinical change and volumetric change, the SPM multiple regression analyses including clinical severity reduction (change in QIDS score) and anhedonia symptomatology improvement (change in SHAPS score) as independent predictors and voxel-wise GM signal changes within the right NAcc, VTA, amygdala, mOFC, hippocampus and BA25 (i.e., using small-volume correction procedures) as dependent variables did not reveal significant findings.

By contrast, when assessing group differences between responders and non-responders, QIDS responders showed a significantly higher post-treatment GM volume increase in the right mOFC, the right BA25, the right amygdala and the right NAcc. No significant QIDS findings were associated with hippocampus or VTA. In addition, SHAPS responders displayed a significantly higher post-treatment GM volume increase in the right hippocampus and the right amygdala compared to SHAPS non-responders. Non-responders also showed volumetric increases in these structures, but significantly smaller (Table 1 and Figure S1). In contrast, while SHAPS responders also exhibited right VTA GM volume increase, non-responders showed right VTA post-treatment GM volume decreases, revealing a significant and qualitatively different volumetric response to ECT between anhedonia responders and non-responders (Table 1 and Figure 2). No significant SHAPS findings were associated with BA25, mOFC or NAcc.

**Table 1.** Brain volumetric increases in clinical responders versus non-responders

|  | x | y | z | *t* value | *p* value<sup>1</sup> | Anatomical location |
| --- | --- | --- | --- | --- | --- | --- |
| Clinical severity | 6 | 21 | -20 | 6.41 | 0.002 | Right mOFC |
|  | 8 | 23 | -17 | 5.36 | 0.003 | Right BA25 |
|  | 27 | 0 | -12 | 4.36 | 0.007 | Right amygdala |
|  | 14 | 3 | -12 | 3.13 | 0.021 | Right NAcc |
| Anhedonic symptomatology | 33 | -35 | -11 | 5.25 | 0.008 | Right hippocampus |
|  | 35 | 0 | -20 | 3.64 | 0.021 | Right amygdala |
|  | 2 | -21 | -18 | 3.61 | 0.015 | Right VTA |

*mOFC*, medial orbitofrontal cortex; *BA25*, Brodmann area 25; *NAcc*, nucleus accumbens; *VTA*, ventral tegmental area.
*x*, *y*, *z* coordinates are reported in standard Montreal Neurological Institute (MNI) space.
<sup>1</sup> FWE corrected for multiple comparisons (i.e., number of in-mask voxels) and significant after FDR correction for multiple comparisons (i.e., number of masks).

**Figure 2.**
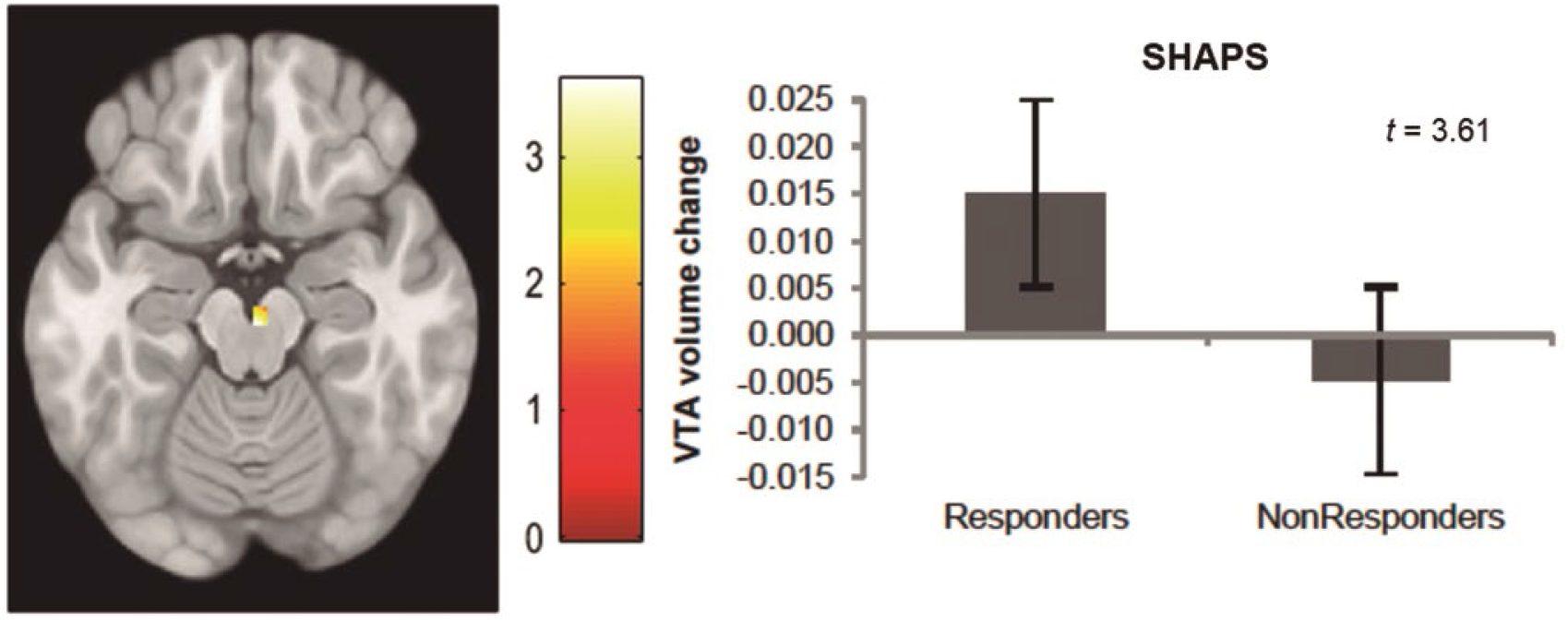
*Left*: Volume increases in SHAPS responders vs. non-responders located at the right ventral tegmental area (VTA). Left hemisphere is depicted on the left. Color bar represents *t*-value. *Right*: Bar plot depicting right VTA gray matter volume changes (peak values) in SHAPS responders and non-responders. Error bars display the standard error.

### Relationship with anticipatory and consummatory subconstructs

The multiple regression analysis including the anticipatory subconstruct of reward processing (i.e., change in anticipatory TEPS score) as the independent predictor showed a significant positive association between change in the anticipation of pleasure capability and regional GM volume increases in the right VTA, right NAcc, right hippocampus and right amygdala (see Table 2 and Figure 3). No significant findings were associated with BA25 or mOFC. Also, no significant findings were observed for the consummatory component of the hedonic experience.

**Table 2.** Brain volumetric increases related to anticipatory pleasure

|  | **x** | **y** | **z** | **t value** | **p value<sup>1</sup>** | **Anatomical location** |
| --- | --- | --- | --- | --- | --- | --- |
| Anticipatory pleasure | 30 | -14 | -12 | 5.66 | 0.004 | Right hippocampus |
|  | 26 | -9 | -15 | 4.66 | 0.004 | Right amygdala |
|  | 2 | -17 | -14 | 3.08 | 0.026 | Right VTA |
|  | 9 | 11 | -14 | 2.87 | 0.029 | Right NAcc |

*VTA*, ventral tegmental area; *NAcc*, nucleus accumbens.
*x*, *y*, *z* coordinates are reported in standard Montreal Neurological Institute (MNI) space.
<sup>1</sup> FWE corrected for multiple comparisons (i.e., number of in-mask voxels) and significant after FDR correction for multiple comparisons (i.e., number of masks).

**Figure 3.**
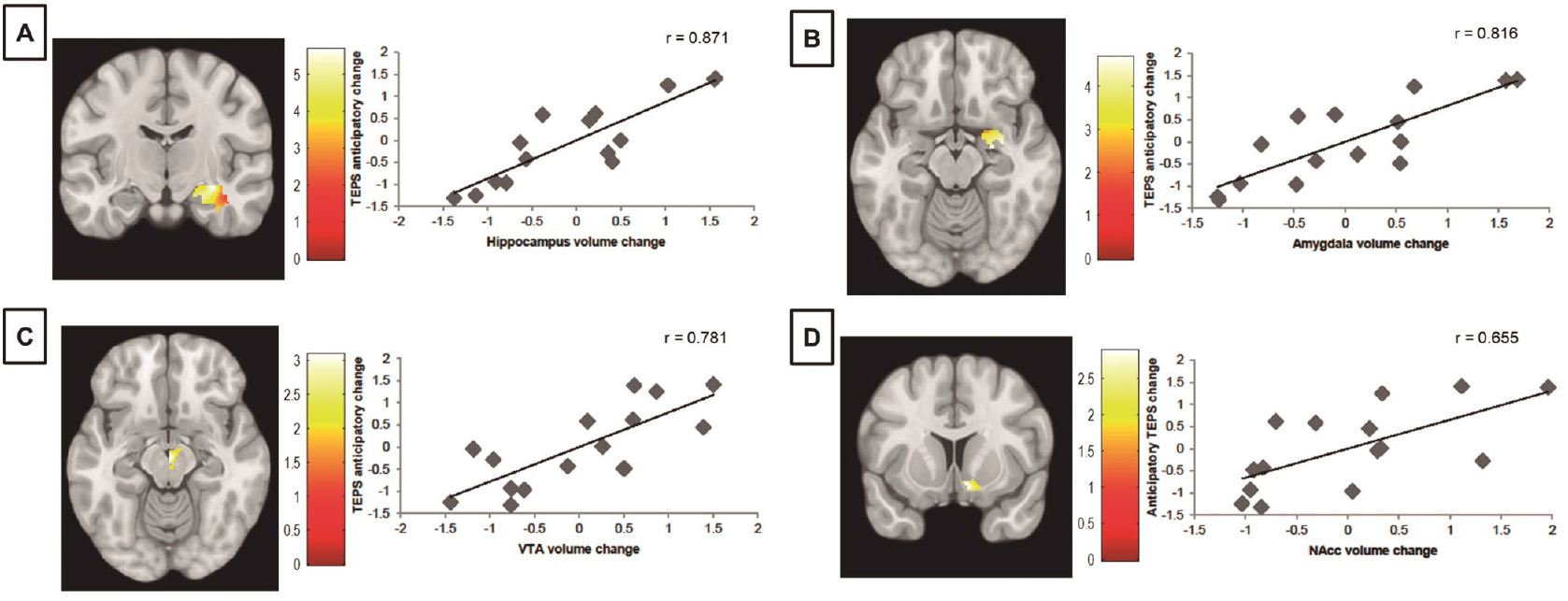
*Left figures*: Volume increases related to anticipatory pleasure improvement (percentage of anticipatory TEPS scores change) located at the right hippocampus (A), right amygdala (B), right ventral tegmental area (VTA, C), and right nucleus accumbens (NAcc, D). Left hemisphere is depicted on the left. Color bar represents *t*-value. *Right figures*: Scatter plots depicting the relationship between regional gray matter volume change (peak values) in the right hippocampus (A), right amygdala (B), right VTA (C), and right NAcc (D) and anticipatory pleasure improvement (percentage of anticipatory TEPS scores change), controlled by age and gender.

## DISCUSSION

In the present study we assessed the specific impact of ECT in ameliorating maladaptive reward processing in patients with TRD, observing a significant improvement not only in overall syndromal depression severity (QIDS), but also in anhedonia (SHAPS) and both anticipatory and consummatory dimensional reward constructs (anticipatory and consummatory TEPS) after RUL ECT. Anatomically, we detected a significant effect separating anhedonia (SHAPS) responders and non-responders in ECT-induced GM volume increases within the right VTA, right hippocampus and right amygdala (but not within the right BA25, mOFC and NAcc). Dimensionally, we observed a significant association between the improvement in anticipatory pleasure (but not consummatory pleasure) and GM volume increases in the right VTA, right NAcc, right hippocampus and right amygdala (but not in the right BA25 and mOFC). As a clinical/behavioral control in order to evaluate the specificity of these effects on reward constructs, we also assessed the impact of ECT-induced GM volume increases on overall depression severity (QIDS). We found a significant effect between responders and non-responders in the right mOFC, the right BA25, the right amygdala and the right NAcc (but not in the right hippocampus and right VTA). Notwithstanding, we should note the lack of significant correlations when evaluating depression severity and anhedonia symptoms as continuum variables (as opposed to categoric responders and non-responders analyses).

The term anhedonia was first defined by Ribot [31] as the inability to experience pleasure. Later, Meehl [32] introduced the “hedonic capacity model of anhedonia”, which suggested that anhedonic patients show deficits in the subjective effectiveness of positive reinforcers. Although this theory suggested that the anticipation of a future reward is promoted by the pleasure of the reward itself, Klein [33] shortly afterward specifically highlighted the distinction between anticipatory and consummatory pleasure. He theorized that anticipatory pleasure was more closely linked to motivation and goal-directed behavior, whereas consummatory pleasure was more closely linked to satiation. Moreover, previous studies in depressed patients with high levels of anhedonia also support the uncoupling between anticipatory and consummatory pleasure. Hanley [34], for instance, revealed that patients with depression showed a deficient ability to anticipate pleasure, while their reports of consummatory pleasure did not differ from healthy participants. Similarly, Sherdell et al., [35] noticed that deficits in motivation for reward in patients with depression were primarily driven by low anticipatory pleasure but not decreased consummatory liking. Interestingly, our findings reveal that ECT-related improvements in anticipatory pleasure are significantly greater than consummatory pleasure response. In this sense, a better understanding of the dimensional maladaptive mechanisms underlying the different subdomains of anhedonia is needed to formulate better pathophysiological models of mood disorders, particularly at the circuit level, which are critical to support translational research, define severity, predictor and response biomarkers, and develop novel treatments.

Several brain regions within the cortico-basal ganglia network are part of the reward circuit [25]. However, the existence of convergent reward-related innervations within the NAcc and the VTA, their structural connectivity via the medial forebrain bundle (or mesolimbic pathway), and their common modulation by dopamine (DA), places these regions as key brain structures for reward processing [23,24]. In this sense, our findings highlight the VTA volume as an anhedonia response biomarker to ECT: not only did anhedonic responders show a GM volume increase in the VTA, but non-responders displayed a GM volume decrease within the same brain region. More specifically, Berridge and Robinson [36] observed that a discrete manipulation of the VTA significantly altered anticipatory reward functions. Likewise, Berridge and Robinson [37] acknowledged that the NAcc consummatory hedonic hotspot constitutes only 10% of total NAcc volume whereas the remaining 90% is responsible for anticipatory pleasure. These reports are in agreement with our findings revealing that NAcc-VTA pathway is closely related to ECT-induced anticipatory response (Figure 3) but not consummatory response.

Moreover, Sesack and Grace [24] highlighted that the hippocampus is one of the best-positioned brain regions to provide a modulatory gating influence over the NAcc and the VTA. Indeed, the hippocampus-NAcc modulation seems to be responsible for the spontaneous firing state of DA neurons in the VTA [38,39], and, interestingly, Gauthier and Tank [40] recently identified a population of reward-associated neurons in the hippocampus. In this sense, reward-related stimuli appear to trigger a hippocampus-NAcc-VTA drive potentiation to reinforce ongoing behavior [25]. Less reliable evidence supports amygdalar activity in contexts involving potential reward than in those involving potential punishment [41]. Indeed, data from animal literature and human neuroimaging studies have delineated the amygdala as a critical brain structure for fear conditioning and negative valence affective processing [42]. Nonetheless, Baxter and Murray [43] suggested that although reward processing has not been typically associated with amygdala function, amygdala contributions to stimulus-reward learning should be expected (i.e., current stimulus-value associations). To this point, however, it should be noted that GM volume increases in the amygdala and the NAcc were related to both anhedonia and overall depression severity reduction (QIDS score) in our responder analyses. Therefore, our findings as a whole appear to support that the amygdala and NAcc may engage in more complex emotional processes able to shape both specific reward and more global syndromic depression improvement, while a hippocampal-NAcc-VTA loop seems more particular for anticipatory reward responses.

In contrast with most studies being unable to correlate ECT-induced GM volume increases with the improvement in depression severity [44–54], our responder analyses also revealed that depressive responders exhibited higher mOFC and BA25 GM volume increases than non-responders (although non-responders also showed smaller GM volume increases within these brain regions). Unexpectedly, ECT-induced mOFC increases appeared to be more associated with overall syndromal depression severity than with anhedonia, as we hypothesized given the role of the mOFC as a critical cortical hub of the reward network. That said, our anatomical control BA25 was only associated to depression severity response but not to anhedonia or reward processing, as we hypothesized, further highlighting the specificity of the observed ECT-induced effects in reward circuitry anatomy and clinical dimensions. Interestingly, both mOFC and BA25 have been involved in antidepressant treatment response with both pharmacological and invasive neuromodulation therapies [27,55]. Overall, our findings are in agreement with the psychological and neurobiological dissociation between anticipation and consumption of reward, as well as with the critical involvement of anticipatory pleasure in patients with depression. More broadly, we highlight the importance of a dimensional and circuit-based approach to study neuropsychiatric pathophysiology and target engagement. Future research should evaluate the neurobiological correlates of particular clinical dimensions of depression in order to detect symptom-specific response biomarkers for ECT and other device neuromodulation therapies.

This study has a number of limitations. First, we evaluated a relatively modest sample size, which is in line with other single site neuroimaging studies of patients receiving ECT given the complexity of data acquisition in this population. Second, the absence of matched comparison control group; however, the longitudinal design of the study permitted conducting powerful within-subject analyses, with each participant being their own control. Also, we used BA25 as an anatomical control (relevant for depression and mood, but not for reward specifically), and the QIDS as a clinical/behavioral control to understand the reward specificity of our findings. Third, we cannot determine what effect, if any, the concurrent pharmacological treatment had on our results, although in an attempt to minimize this confounding effect, pharmacological treatment was not modified throughout the entire ECT course. Finally, reward system function was evaluated by means of a subjective psychometric assessment. Future studies should attempt to obtain objective evaluations of reward system function by means of validated neuropsychological protocols, ideally, in combination with functional neuroimaging assessments.

In conclusion, our findings highlight the unmet relevant need of transitioning from a syndromical to a dimensional and circuit-level approach to deeply understand neuromodulatory treatment response. Future research should further evaluate structure-symptom dimension relationships to capture complex psychopathological processes aiming to identify useful response biomarkers to support and inform treatment development.

## Data Availability

Statistical maps will be available on demand

## Funding and Disclosure

This study was supported by the NIMH (R01 MH112737-01) and the Harvard Medical School Dupont-Warren award to JAC. MC is funded by a Sara Borrell postdoctoral contract (CD20/00189). JAC is a member of the scientific advisory board of Feel more Labs and Hyka Therapeutics. The rest of authors declared no competing interest to disclose.

## Author contributions

**MC**: Methodology, Formal analysis, Data curation, Writing – original draft, Visualization, Writing – review & editing. **EL**: Investigation, Data curation, Writing – review & editing. **AW**: Investigation, Data curation, Writing – review & editing. **KE**: Conceptualization, Project administration, Resources, Writing – review & editing. **TB**: Conceptualization, Project administration, Resources, Writing – review & editing. **CS-M**: Methodology, Supervision, Writing – review & editing. **JAC**: Conceptualization, Funding Acquisition, Project administration, Resources, Methodology, Supervision, Writing – review & editing.

## Data availability

Statistical maps will be available on demand.

## Notes

### Competing Interest Statement

Dr. Camprodon is a member of the scientific advisory board of Feel more Labs and Hyka Therapeutics. The rest of authors declared no competing interest to disclose.

### Funding Statement

This study was supported by the NIMH (R01 MH112737-01) and the Harvard Medical School Dupont-Warren award to Dr. Camprodon. Dr. Cano is funded by a Sara Borrell postdoctoral contract (CD20/00189).

### Author Declarations

Ethics committee/IRB of Massachusetts General Hospital gave ethical approval for this work

## References

1. Jaffe R. The Practice of Electroconvulsive Therapy: Recommendations for Treatment, Training, and Privileging: A Task Force Report of the American Psychiatric Association, 2nd ed. Am J Psychiatry. 2002;159:331–331.

2. Tor PC, Tan XW, Martin D, Loo C. Comparative outcomes in electroconvulsive therapy (ECT): A naturalistic comparison between outcomes in psychosis, mania, depression, psychotic depression and catatonia. Eur Neuropsychopharmacol. 2021;51:43–54.

3. Fink M. What was learned: studies by the consortium for research in ECT (CORE) 1997-2011. Acta Psychiatr Scand. 2014;129:417–426.

4. Lee WH, Deng Z-D, Kim T-S, Laine AF, Lisanby SH, Peterchev AV. Regional electric field induced by electroconvulsive therapy in a realistic finite element head model: influence of white matter anisotropic conductivity. NeuroImage. 2012;59:2110–2123.

5. Ottosson JO. Experimental studies of the mode of action of electroconvulsive therapy: Introduction. Acta Psychiatr Scand Suppl. 1960;35:5–6.

6. Abrams R. Seizure Generalization and Unilateral Electroconvulsive Therapy. Convuls Ther. 1991;7:213–217.

7. Cano M, Martínez-Zalacaín I, Bernabéu-Sanz Á, Contreras-Rodríguez O, Hernández-Ribas R, Via E, et al. Brain volumetric and metabolic correlates of electroconvulsive therapy for treatment-resistant depression: a longitudinal neuroimaging study. Transl Psychiatry. 2017;7:e1023.

8. Cano M, Lee E, Cardoner N, Martínez-Zalacaín I, Pujol J, Makris N, et al. Brain volumetric correlates of right unilateral versus bitemporal electroconvulsive therapy for treatment-resistant depression. J Neuropsychiatry Clin Neurosci. 2019;31:152–158.

9. Whitton AE, Treadway MT, Pizzagalli DA. Reward processing dysfunction in major depression, bipolar disorder and schizophrenia. Curr Opin Psychiatry. 2015;28:7–12.

10. Sackeim HA, Prudic J, Fuller R, Keilp J, Lavori PW, Olfson M. The cognitive effects of electroconvulsive therapy in community settings. Neuropsychopharmacol Off Publ Am Coll Neuropsychopharmacol. 2007;32:244–254.

11. Semkovska M, McLoughlin DM. Objective cognitive performance associated with electroconvulsive therapy for depression: a systematic review and meta-analysis. Biol Psychiatry. 2010;68:568–577.

12. Martin DM, McClintock SM, Loo CK. Brief cognitive screening instruments for electroconvulsive therapy: Which one should I use? Aust N Z J Psychiatry. 2020;54:867–873.

13. Landry M, Moreno A, Patry S, Potvin S, Lemasson M. Current Practices of Electroconvulsive Therapy in Mental Disorders: A Systematic Review and Meta-Analysis of Short and Long-Term Cognitive Effects. J ECT. 2021;37:119–127.

14. Insel T, Cuthbert B, Garvey M, Heinssen R, Pine DS, Quinn K, et al. Research domain criteria (RDoC): toward a new classification framework for research on mental disorders. Am J Psychiatry. 2010;167:748–751.

15. Rømer Thomsen K, Whybrow PC, Kringelbach ML. Reconceptualizing anhedonia: novel perspectives on balancing the pleasure networks in the human brain. Front Behav Neurosci. 2015;9:49.

16. Treadway MT, Zald DH. Reconsidering anhedonia in depression: lessons from translational neuroscience. Neurosci Biobehav Rev. 2011;35:537–555.

17. Gard DE, Kring AM, Gard MG, Horan WP, Green MF. Anhedonia in schizophrenia: distinctions between anticipatory and consummatory pleasure. Schizophr Res. 2007;93:253–260.

18. Sheehan DV, Lecrubier Y, Sheehan KH, Amorim P, Janavs J, Weiller E, et al. The Mini-International Neuropsychiatric Interview (M.I.N.I.): the development and validation of a structured diagnostic psychiatric interview for DSM-IV and ICD-10. J Clin Psychiatry. 1998;59 Suppl 20:22–33;quiz 34-57.

19. Rush AJ, Trivedi MH, Ibrahim HM, Carmody TJ, Arnow B, Klein DN, et al. The 16-Item Quick Inventory of Depressive Symptomatology (QIDS), clinician rating (QIDS-C), and self-report (QIDS-SR): a psychometric evaluation in patients with chronic major depression. Biol Psychiatry. 2003;54:573–583.

20. Snaith RP, Hamilton M, Morley S, Humayan A, Hargreaves D, Trigwell P. A scale for the assessment of hedonic tone the Snaith-Hamilton Pleasure Scale. Br J Psychiatry J Ment Sci. 1995;167:99–103.

21. Gard DE, Gard MG, Kring AM, John OP. Anticipatory and consummatory components of the experience of pleasure: A scale development study. J Res Personal. 2006;40:1086–1102.

22. d’Elia G. Unilateral electroconvulsive therapy. Acta Psychiatr Scand Suppl. 1970;215:1–98.

23. Russo SJ, Nestler EJ. The brain reward circuitry in mood disorders. Nat Rev Neurosci. 2013;14:609–625.

24. Sesack SR, Grace AA. Cortico-Basal Ganglia reward network: microcircuitry. Neuropsychopharmacol Off Publ Am Coll Neuropsychopharmacol. 2010;35:27–47.

25. Haber SN, Knutson B. The reward circuit: linking primate anatomy and human imaging. Neuropsychopharmacol Off Publ Am Coll Neuropsychopharmacol. 2010;35:4–26.

26. Berridge KC, Robinson TE. Parsing reward. Trends Neurosci. 2003;26:507–513.

27. Seminowicz DA, Mayberg HS, McIntosh AR, Goldapple K, Kennedy S, Segal Z, et al. Limbic-frontal circuitry in major depression: a path modeling metanalysis. NeuroImage. 2004;22:409–418.

28. Maldjian JA, Laurienti PJ, Kraft RA, Burdette JH. An automated method for neuroanatomic and cytoarchitectonic atlas-based interrogation of fMRI data sets. NeuroImage. 2003;19:1233–1239.

29. Murty VP, Shermohammed M, Smith DV, Carter RM, Huettel SA, Adcock RA. Resting state networks distinguish human ventral tegmental area from substantia nigra. NeuroImage. 2014;100:580–589.

30. Benjamini Y, Hochberg Y. Controlling the False Discovery Rate: A Practical and Powerful Approach to Multiple Testing. J R Stat Soc Ser B Methodol. 1995;57:289–300.

31. Ribot T. The Psychology of Emotions. London, UK: W. Scott; 1897.

32. Meehl PE. Hedonic capacity: some conjectures. Bull Menninger Clin. 1975;39:295–307.

33. Klein D. Depression and anhedonia. In: Clark C, Fawcett J, editors. Anhedonia and Affect Deficits States. New York, NY: PMA Publishing; 1984.

34. Hanley K. Anhedonia and depression: Anticipatory, Consummatory, and Recall Deficits. 2007. https://digitalcommons.colby.edu/cgi/viewcontent.cgi?article=1113&context=honorstheses. Accessed 25 October 2021.

35. Sherdell L, Waugh CE, Gotlib IH. Anticipatory pleasure predicts motivation for reward in major depression. J Abnorm Psychol. 2012;121:51–60.

36. Berridge KC, Robinson TE. What is the role of dopamine in reward: hedonic impact, reward learning, or incentive salience? Brain Res Brain Res Rev. 1998;28:309–369.

37. Berridge KC, Robinson TE. Liking, Wanting and the Incentive-Sensitization Theory of Addiction. Am Psychol. 2016;71:670–679.

38. Lisman JE, Grace AA. The hippocampal-VTA loop: controlling the entry of information into long-term memory. Neuron. 2005;46:703–713.

39. Floresco SB, Todd CL, Grace AA. Glutamatergic Afferents from the Hippocampus to the Nucleus Accumbens Regulate Activity of Ventral Tegmental Area Dopamine Neurons. J Neurosci. 2001;21:4915–4922.

40. Gauthier JL, Tank DW. A Dedicated Population for Reward Coding in the Hippocampus. Neuron. 2018;99:179–193.e7.

41. Zald DH. The human amygdala and the emotional evaluation of sensory stimuli. Brain Res Brain Res Rev. 2003;41:88–123.

42. Milad MR, Quirk GJ. Fear extinction as a model for translational neuroscience: ten years of progress. Annu Rev Psychol. 2012;63:129–151.

43. Baxter MG, Murray EA. The amygdala and reward. Nat Rev Neurosci. 2002;3:563–573.

44. Nordanskog P, Dahlstrand U, Larsson MR, Larsson E-M, Knutsson L, Johanson A. Increase in hippocampal volume after electroconvulsive therapy in patients with depression: a volumetric magnetic resonance imaging study. J ECT. 2010;26:62–67.

45. Nordanskog P, Larsson MR, Larsson E-M, Johanson A. Hippocampal volume in relation to clinical and cognitive outcome after electroconvulsive therapy in depression. Acta Psychiatr Scand. 2014;129:303–311.

46. Tendolkar I, van Beek M, van Oostrom I, Mulder M, Janzing J, Voshaar RO, et al. Electroconvulsive therapy increases hippocampal and amygdala volume in therapy refractory depression: a longitudinal pilot study. Psychiatry Res. 2013;214:197–203.

47. Abbott CC, Jones T, Lemke NT, Gallegos P, McClintock SM, Mayer AR, et al. Hippocampal structural and functional changes associated with electroconvulsive therapy response. Transl Psychiatry. 2014;4:e483.

48. Ota M, Noda T, Sato N, Okazaki M, Ishikawa M, Hattori K, et al. Effect of electroconvulsive therapy on gray matter volume in major depressive disorder. J Affect Disord. 2015;186:186–191.

49. Bouckaert F, De Winter F-L, Emsell L, Dols A, Rhebergen D, Wampers M, et al. Grey matter volume increase following electroconvulsive therapy in patients with late life depression: a longitudinal MRI study. J Psychiatry Neurosci JPN. 2016;41:105–114.

50. Jorgensen A, Magnusson P, Hanson LG, Kirkegaard T, Benveniste H, Lee H, et al. Regional brain volumes, diffusivity, and metabolite changes after electroconvulsive therapy for severe depression. Acta Psychiatr Scand. 2016;133:154–164.

51. Sartorius A, Demirakca T, Böhringer A, Clemm von Hohenberg C, Aksay SS, Bumb JM, et al. Electroconvulsive therapy increases temporal gray matter volume and cortical thickness. Eur Neuropsychopharmacol J Eur Coll Neuropsychopharmacol. 2016;26:506–517.

52. Bouckaert F, Dols A, Emsell L, De Winter F-L, Vansteelandt K, Claes L, et al. Relationship Between Hippocampal Volume, Serum BDNF, and Depression Severity Following Electroconvulsive Therapy in Late-Life Depression. Neuropsychopharmacol Off Publ Am Coll Neuropsychopharmacol. 2016;41:2741–2748.

53. Qiu H, Li X, Zhao W, Du L, Huang P, Fu Y, et al. Electroconvulsive Therapy-Induced Brain Structural and Functional Changes in Major Depressive Disorders: A Longitudinal Study. Med Sci Monit Int Med J Exp Clin Res. 2016;22:4577–4586.

54. Oltedal L, Narr KL, Abbott C, Anand A, Argyelan M, Bartsch H, et al. Volume of the Human Hippocampus and Clinical Response Following Electroconvulsive Therapy. Biol Psychiatry. 2018;84:574–581.

55. Mayberg HS, Lozano AM, Voon V, McNeely HE, Seminowicz D, Hamani C, et al. Deep Brain Stimulation for Treatment-Resistant Depression. Neuron. 2005;45:651–660.

