## Supplementary Material for "Electroconvulsive therapy effects on anhedonia and reward circuitry anatomy: a dimensional structural neuroimaging approach"

**Table S1.** Sociodemographic, clinical and reward-related characteristics of the study sample

| *Sociodemographic, clinical and reward-related variables* | **TRD patients (n=15)** |
|---|---|
| **Age**, years: mean (s.d.) | 42.93 (14.87) |
| **Gender**, male: n (%) | 7 (46.67) |
| **Drugs:** % | |
| Antidepressant | 93.33 |
| Antipsychotics | 40 |
| Anxiolytics | 46.66 |
| **QIDS1**[1]: mean (s.d.) | 18 (3.38) |
| **QIDS2**: mean (s.d.) | 12 (5.21) |
| **QIDS change**: mean (s.d.); % | 6 (6.5); 30.40 |
| **QIDS response rate**: n (%) | 5 (33.34) |
| **SHAPS[1]**: mean (s.d.) | 5.07 (2.76) |
| **SHAPS[2]**: mean (s.d.) | 2.47 (3.31) |
| **SHAPS change**: mean (s.d.); % | 2.6 (2.87); 52.16 |
| **SHAPS response rate**: n (%) | 10 (66.67) |
| **TEPS1**[1]**:** mean (s.d.) | |
| Anticipatory score | 41 (9.91) |
| Consummatory score | 42.20 (9.99) |
| **TEPS2:** mean (s.d.) | |
| Anticipatory score | 50.47 (9.02) |
| Consummatory score | 45.67 (9.19) |
| **TEPS change**: mean (s.d.); % | |
| Anticipatory score | 9.47 (8.62); 17.96 |
| Consummatory score | 3.47 (4.40); 7.88 |

*TRD*, Treatment-Resistant Depression

[1]This number indicates the time-point corresponding to the Quick Inventory of Depressive Symptomatology (QIDS), the Snaith-Hamilton Pleasure Scale (SHAPS) and the Temporal Experience of Pleasure Scale (TEPS) scores. (1) Score in the 1$^{st}$ neuroimaging assessment and (2) Score in the 2$^{nd}$ neuroimaging assessment.

**Table S2.** Brain areas showing gray matter volume increases in patients with treatment-resistant depression treated with right unilateral electroconvulsive therapy

| x | y | z | *t* value | *p* value[1] | Anatomical location |
|---|---|---|---|---|---|
| 21 | -2 | -9 | 8.49 | 0.009 | Right pallidum |
| 27 | -12 | -11 | 7.52 | 0.023 | Right hippocampus |
| 9 | 21 | -9 | 7.36 | 0.027 | Right SgACC |
| 54 | 3 | -2 | 7.02 | 0.039 | Right superior temporal gryus |

*SgACC*, subgenual anterior cingulate cortex.
*x, y, z* coordinates are reported in standard Montreal Neurological Institute (MNI) space.
[1] FWE corrected for multiple comparisons.

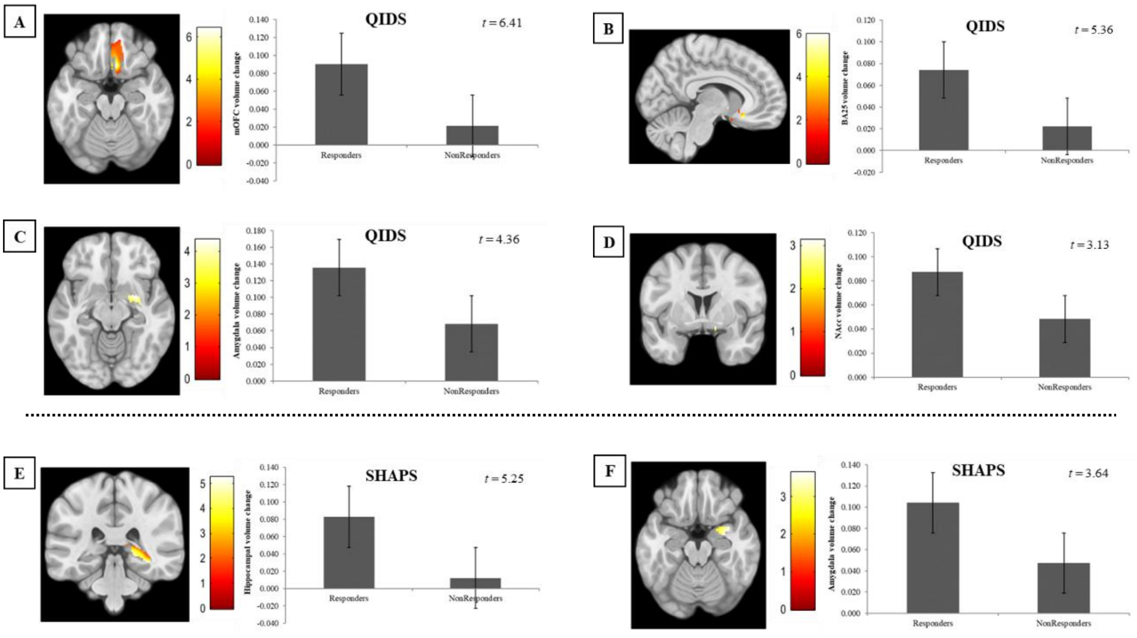


**Figure S1.** *Left figures*: Volume increases in QIDS responders vs. non-responders located at the right medial orbitofrontal cortex (mOFC, A), right Brodmann area 25 (BA25, B), right amygdala (C), and right nucleus accumbens (NAcc, D). Volume increases in SHAPS responders vs. non-responders located at the right hippocampus (E), and right amygdala (F). Left hemisphere is depicted on the left. Color bar represents *t*-value. *Right figures*: Bar plots depicting right mOFC (A), right BA25 (B), right amygdala (C), and right NAcc (D) gray matter volume changes (peak values) in QIDS responders and non-responders. Bar plots depicting right hippocampus (E), and right amygdala (F) gray matter volume changes (peak values) in SHAPS responders and non-responders. Error bars display the standard error.
